# High Extracellular-to-Intracellular Water Ratio in Limb Muscles is Associated with Low Muscle Strength in Patients with Heart Failure

**DOI:** 10.1101/2023.08.09.23293909

**Authors:** Kensuke Nakamura, Yoshiharu Kinugasa, Takeshi Sota, Masayuki Hirai, Masahiko Kato, Kazuhiro Yamamoto

## Abstract

**Background:** A high extracellular water (ECW) to intracellular water (ICW) ratio of skeletal muscle as assessed by bioelectrical impedance analysis is reportedly associated with loss of muscle strength. However, the validity of this index for heart failure (HF), which is likely associated with changes in the water distribution, is unclear.

**Methods:** This study involved 190 patients with HF. The total ECW and ICW of both upper and lower extremities were measured, and a high ECW/ICW ratio was defined as an ECW/ICW ratio higher than the median (≥0.636 for men, ≥0.652 for women). Low muscle strength was defined as reduced handgrip strength according to the criteria established by the Asian Working Group for Sarcopenia.

**Results:** Patients with a high ECW/ICW ratio were older, had a higher left ventricular ejection fraction and B-type natriuretic peptide level, and had a lower body mass index, hemoglobin level, albumin level, estimated glomerular filtration rate, handgrip strength, and 6-minute walk distance than patients with a low ECW/ICW ratio (p < 0.05). An increasing ECW and/or decreasing ICW was associated with a higher ECW/ICW ratio (p < 0.05). In the multivariate logistic regression analysis, a high ECW/ICW ratio and low skeletal muscle mass were independently associated with low muscle strength (p < 0.05).

**Conclusion:** A high ECW/ICW ratio in limb muscles (i.e., an increasing ECW and/or decreasing ICW) is independently associated with muscle weakness regardless of skeletal muscle mass in patients with HF.

*What Is New?:*

◆ A high extracellular water (ECW) to intracellular water (ICW) ratio of skeletal muscle as assessed by bioelectrical impedance analysis was independently associated with low muscle strength regardless of skeletal muscle mass in patients with heart failure (HF).
◆ Both an increasing ECW and decreasing ICW were independently associated with low muscle strength in patients with HF.

*What Are the Clinical Implications?:*

◆ The ECW/ICW ratio in limb muscles is a new index that can be used to assess muscle function apart from muscle mass in patients with HF.

## Introduction

The number of older patients with heart failure (HF) is rapidly increasing worldwide. Physical frailty is a common condition in this population, and it leads to worse quality of life and outcomes such as impaired daily function, falls, hospitalization, and mortality.^1^ Physical frailty is largely caused by skeletal muscle dysfunction, which results from changes in both the quantity and quality of skeletal muscle.^2^ Patients with HF have structural and functional abnormalities in their skeletal muscle, such as altered myofiber types, increased fat infiltration, mitochondrial dysfunction, decreased capillary density, and impaired oxygen and metabolite transport.^3^ Research has demonstrated the usefulness of noninvasive assessment of structural and functional changes in skeletal muscle using computed tomography,^4^ magnetic resonance imaging,^5^ ultrasound,^6^ and bioelectrical impedance analysis (BIA).^7^ BIA can be used to measure the impedance of body parts by applying a weak electric current; it can also enable body composition analysis. Moreover, BIA can be used to measure the intracellular water (ICW) and extracellular water (ECW) of skeletal muscle by using multiple frequencies. ICW reflects the water volume of muscle cells, and ECW reflects the water volume of extracellular spaces.^8,9^ A high ECW/ICW ratio has been associated with muscle weakness in patients with chronic kidney disease, patients with liver disease, and healthy older people.^10–12^ However, the validity of the ECW/ICW ratio in patients with HF, which is likely associated with changes in the water distribution, is unclear. The present study was performed to examine the relationship between the ECW/ICW ratio and muscle strength in patients with HF.

## Methods

### Patients

In this retrospective observational study, we screened 976 patients who were hospitalized for acute decompensated HF without pulmonary embolism or acute myocardial infarction and discharged from Tottori University Hospital from April 2014 to March 2022. We excluded 352 patients with missing body composition data and 434 patients with missing handgrip strength data. Finally, we analyzed 190 patients.

We defined HF as the presence of symptoms and signs of HF according to the Framingham criteria, along with pulmonary congestion on X-rays and/or elevated left atrial pressure on Doppler echocardiography as previously described.^13^ All patients received adequate decongestion therapy during hospitalization. After their HF had been stabilized, the patients underwent cardiac rehabilitation during their hospital stay.

### Data collection

We reviewed the patients’ medical records to gather data on their demographics, medical history, comorbidities, laboratory data, echocardiograms, medications, body composition, and physical function test results. We obtained all measurements at discharge when HF was stable. We classified the etiology of HF as ischemic or non-ischemic heart disease. Ischemic heart disease was defined as the presence of significant coronary artery disease and/or a history of coronary artery intervention. We measured the patients’ echocardiographic data according to the recommendations of the American Society of Echocardiography.^14^

### Measurement of body composition

Body composition was measured using a BIA device (InBody 770; InBody Co., Ltd., Seoul, South Korea) as previously described.^13^ The patients stood barefoot on the foot electrodes of the device, their weight equally distributed on both feet in a completely vertical position, and held the hand electrodes with both hands. Thirty impedance measurements were taken at five body sites (right and left arms, trunk, and right and left legs) using six different frequencies (1, 5, 50, 250, 500, and 1000 kHz).^15^

The skeletal muscle mass of the upper and lower extremities was measured, and the skeletal muscle mass index (SMI) was calculated as follows^13^: SMI = extremity skeletal muscle mass / (height)^2^. ECW and ICW of the upper and lower extremities were measured and similarly divided by the square of the height to correct for body size.^16^

### Physical performance

Handgrip strength was measured with a grip dynamometer (GRIP-D; Takei Scientific Instruments Co., Ltd., Niigata, Japan) in the standing position with the upper limb abducted about 20 degrees from the body.^13^ Each hand was measured twice, and the maximum value was used as the representative value. The 6-minute walk distance was measured as previously described^13^ and was available for 90 of the study patients.

### Definition of high ECW/ICW ratio, low skeletal muscle mass, and low muscle strength

A high ECW/ICW ratio was defined as an ECW/ICW ratio higher than the median for each sex because the ratio differed between men and women: ≥0.636 for men and ≥0.652 for women. Low muscle mass and low muscle strength were defined according to the criteria established by the Asian Working Group for Sarcopenia.^17^ Low muscle mass was defined as an SMI of <7.0 kg/m^2^ for men and <5.7 kg/m^2^ for women. Low muscle strength was defined as a handgrip strength of <28 kg for men and <18 kg for women.

### Ethical considerations

This investigation conformed to the Declaration of Helsinki principles. The research ethics committee of Tottori University approved the study. Written informed consent from patients was not required because this was a retrospective observational study of medical records. The study was posted on the hospital’s website according to the Japanese Ministry of Health, Labour and Welfare guidelines; patients were informed of their right to refuse participation in the study, and opt-out consent was obtained.

### Statistical analysis

Continuous variables are expressed as mean ± standard deviation for normally distributed variables and median (interquartile range) for non-normally distributed variables. Categorical variables are expressed as percentages. The t-test was used to compare continuous variables with a normal distribution, and the Mann–Whitney U test was used to compare those without a normal distribution. Fisher’s exact test was used to compare categorical variables. The correlation between handgrip strength and the ECW/ICW ratio was tested using Pearson’s correlation analysis. Multivariate logistic regression analysis was used to assess the factors associated with low muscle strength. All clinical variables that were associated with low muscle strength in the univariate analysis were entered into the multivariate analysis. The following analyses of the combined effect of two indicators were performed using the thin plate smoothing spline: the combined effect of ECW and ICW on the ECW/ICW ratio, the combined effect of ECW and ICW on handgrip strength, and the combined effect of the ECW/ICW ratio and SMI on handgrip strength. These spline models were adjusted for sex because there were sex-related differences in muscle strength and the ECW/ICW ratio. A p value of <0.05 was considered statistically significant. All analyses were performed using R version 4.1.2 (R Foundation for Statistical Computing, Vienna, Austria).

## Results

### Characteristics of patients with low and high ECW/ICW ratio

Table 1 shows the characteristics of the entire study cohort. The median age of the patients was 75 years (60–83), and 64.2% were male. The prevalence of ischemic heart disease and atrial fibrillation was 25.3% and 41.1%, respectively. The mean left ventricular ejection fraction was 48.6% ± 15.5%. A New York Heart Association functional classification of III or IV and a history of HF hospitalization were prevalent in 22.1% and 23.2% of patients, respectively. Non-cardiac comorbidities, specifically chronic obstructive pulmonary disease, diabetes mellitus, and severe renal dysfunction (stage 4 or 5 chronic kidney disease, defined as an estimated glomerular filtration rate of <30 mL/min/1.73 m^2^), were found in 6.3%, 32.8%, and 20.5% of the patients, respectively. The prevalence of loss of muscle mass and loss of muscle strength as defined by the Asian Working Group for Sarcopenia diagnostic criteria was 65.7% and 55.3%, respectively. Low muscle strength was observed in 30.8% of patients with preserved muscle mass. By contrast, 32% of patients with reduced muscle mass had preserved muscle strength.

**Table 1.** Characteristics of patients with low and high ECW/ICW ratio.

| Characteristics | Overall<br>(n=190) | Low ECW/ICW ratio<br>(n=95) | High ECW/ICW ratio<br>(n=95) | P value |
| --- | --- | --- | --- | --- |
| Age (years) | 75 (60-83) | 69 (49-79) | 81 (71-86) | <0.001 |
| Male | 122 (64.2) | 61 (64.2) | 61 (64.2) | 1.000 |
| BMI (kg/m<sup>2</sup>) | 21.6±4.2 | 22.2±4.4 | 20.9±3.9 | 0.029 |
| Systolic blood pressure (mmHg) | 113±20 | 112±18 | 114±22 | 0.460 |
| Prior heart failure admissions | 44 (23.2) | 20 (21.1) | 24 (25.3) | 0.610 |
| LVEF (%) | 48.6±15.5 | 44.9±16.9 | 52.2±13.0 | 0.001 |
| NYHA Class III/IV (%) | 42 (22.1) | 13 (13.7) | 29 (30.5) | 0.008 |
| Ischemic heart disease | 48 (25.3) | 18 (18.9) | 30 (31.6) | 0.066 |
| Co-morbidities |  |  |  |  |
| Hypertension | 125 (65.8) | 55 (57.9) | 70 (73.7) | 0.032 |
| Diabetes | 61 (32.1) | 26 (27.4) | 35 (36.8) | 0.210 |
| Dyslipidemia | 84 (44.2) | 37 (38.9) | 47 (49.5) | 0.188 |
| Atrial fibrillation | 78 (41.1) | 35 (36.8) | 43 (45.3) | 0.300 |
| COPD | 12 (6.3) | 7 (7.4) | 5 (5.3) | 0.767 |
| Cerebrovascular disease | 23 (12.1) | 11 (11.6) | 12 (12.6) | 1.000 |
| Laboratory value at discharge |  |  |  |  |
| Sodium (mmol/L) | 139±3 | 138±3 | 139±4 | 0.160 |
| Hemoglobin (g/dL) | 12.5±2.5 | 13.7±2.5 | 11.2±1.9 | <0.001 |
| Albumin (g/dL) | 3.7±0.5 | 3.9±0.4 | 3.5±0.4 | <0.001 |
| eGFR (mL/min/1.73 m<sup>2</sup>) | 46.2 (34.0-63.7) | 58.3 (41.5-69.4) | 40.2 (26.5-50.4) | <0.001 |
| CRP (mg/dL) | 0.17 (0.07-0.50) | 0.22 (0.08-0.56) | 0.14 (0.06-0.43) | 0.232 |
| BNP (pg/mL) (n=189) | 209 (118-396) | 167 (93-309) | 286 (155-502) | <0.001 |
| Medications at discharge |  |  |  |  |
| ACE-I/ARB/ARNI | 135 (71.1) | 66 (69.5) | 69 (72.6) | 0.749 |
| β-blockers | 149 (78.4) | 80 (84.2) | 69 (72.6) | 0.077 |
| Mineralocorticoid receptor antagonists | 86 (46.3) | 57 (60.0) | 29 (30.5) | <0.001 |
| SGLT-2 inhibitors | 5 (2.6) | 5 (5.3) | 0 (0.00) | 0.059 |
| Loop diuretics | 172 (90.4) | 87 (91.6) | 85 (89.5) | 0.805 |
| Tolvaptan | 34 (17.9) | 14 (14.7) | 20 (21.1) | 0.344 |
| Skeletal muscle structure |  |  |  |  |
| SMI (kg/m<sup>2</sup>) | 6.2±1.3 | 6.3±1.3 | 6.0±1.4 | 0.132 |
| ECW (L/m<sup>2</sup>) | 1.86±0.38 | 1.86±0.33 | 1.86±0.44 | 0.956 |
| ICW (L/m<sup>2</sup>) | 2.91±0.62 | 3.03±0.59 | 2.78±0.63 | 0.006 |
| ECW/ICW ratio | 0.64±0.03 | 0.62±0.02 | 0.67±0.02 | <0.001 |
| Physical performance |  |  |  |  |
| Hand grip strength (kg) | 24.4±9.3 | 27.6±9.3 | 21.1±8.1 | <0.001 |
| Low muscle strength | 105 (55.3) | 34 (35.8) | 71 (74.7) | <0.001 |
| 6-minute walk distance (m) (n=90) | 389±127 | 440±114 | 329±116 | <0.001 |

Data given as n (%), mean ± standard deviation, or median (interquartile range). BMI, body mass index; GNRI, geriatric nutritional risk index; LVEF, left ventricular ejection fraction; NYHA, New York Heart Association; COPD, chronic obstructive pulmonary disease; BUN, blood urea nitrogen; eGFR, estimated glomerular filtration rate; CRP, C-reactive protein; BNP, B-type natriuretic peptide; ACE-I, angiotensin-converting enzyme inhibitor; ARB, angiotensin receptor blocker; ARNI, angiotensin receptor–neprilysin inhibitor; SGLT-2, sodium–glucose cotransporter 2; ICW, intracellular water; ECW, extracellular water; SMI, skeletal muscle index.

Figure 1 shows the distribution of the ECW/ICW ratio for the entire study cohort and the sex-related difference. The mean and median ECW/ICW ratio for the entire cohort were 0.643 ± 0.033 and 0.641 (0.622–0.663), respectively. The ECW/ICW ratio was significantly higher in women than in men (0.651 ± 0.025 vs. 0.639 ± 0.036, p < 0.05). The median ECW/ICW ratio was 0.636 (0.614–0.659) for men and 0.652 (0.635–0.667) for women.

**Figure 1.**
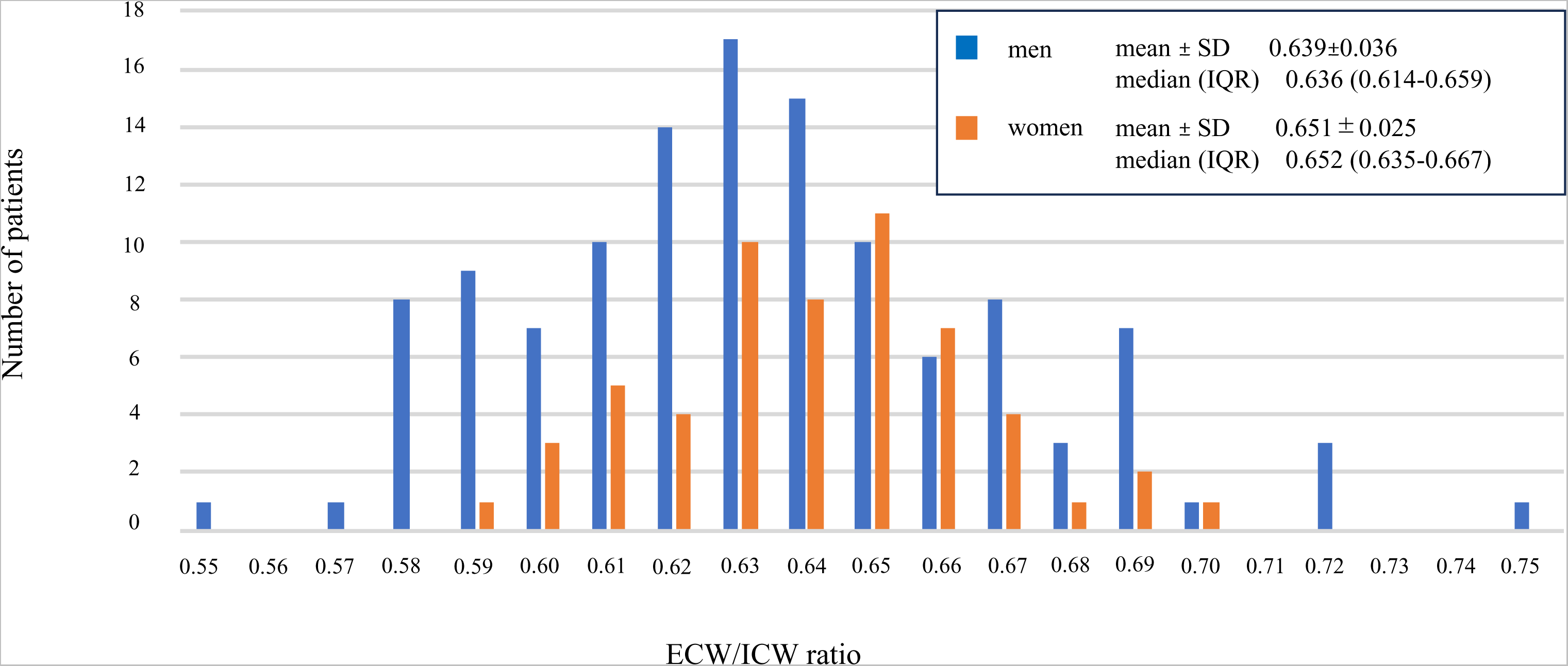
Distribution of the ICW/ ECW ratio in the entire study cohort and sex-related difference in the ratio. ICW, intracellular water; ECW, extracellular water; IQR, interquartile range; SD, standard deviation.

Table 1 shows the characteristics of patients with a low and high ECW/ICW ratio, defined using the median ECW/ICW ratio for each sex. Patients with a high ECW/ICW ratio were older and had a higher incidence of hypertension, higher left ventricular ejection fraction, and higher New York Heart Association functional classification and a lower body mass index than patients with a low ECW/ICW ratio (p < 0.05). The hemoglobin level, albumin level, and estimated glomerular filtration rate were significantly lower and the B-type natriuretic peptide level was significantly higher in patients with a high ECW/ICW ratio than in those with a low ECW/ICW ratio (p < 0.05). Patients with a high ECW/ICW ratio had fewer prescribed mineralocorticoid receptor antagonists than those with a low ECW/ICW ratio (p < 0.05). Most patients’ data were obtained before sodium–glucose transporter 2 inhibitors were allowed to be used for HF in Japan, and the prescription rate of sodium–glucose transporter 2 inhibitors was low in both groups.

### Association of skeletal muscle structure with physical function

Table 1 also shows the association of the ECW/ICW ratio with skeletal muscle structure and physical function. Patients with a high ECW/ICW ratio had a lower ICW, handgrip strength, and 6-minute walk distance than those with a low ECW/ICW ratio (p < 0.05). When analyzed separately for men and women, a high ECW/ICW ratio was associated with decreased handgrip strength in both men and women (32.6 ± 7.0 vs. 24.6 ± 7.4 kg in men and 18.6 ± 5.1 vs. 14.8 ± 4.8 kg in women, respectively; p < 0.05). There was a significant inverse correlation between handgrip strength and the ECW/ICW ratio in both men and women (r = −0.583 and −0.485, respectively; p < 0.05) (Figure 2).

**Figure 2.**
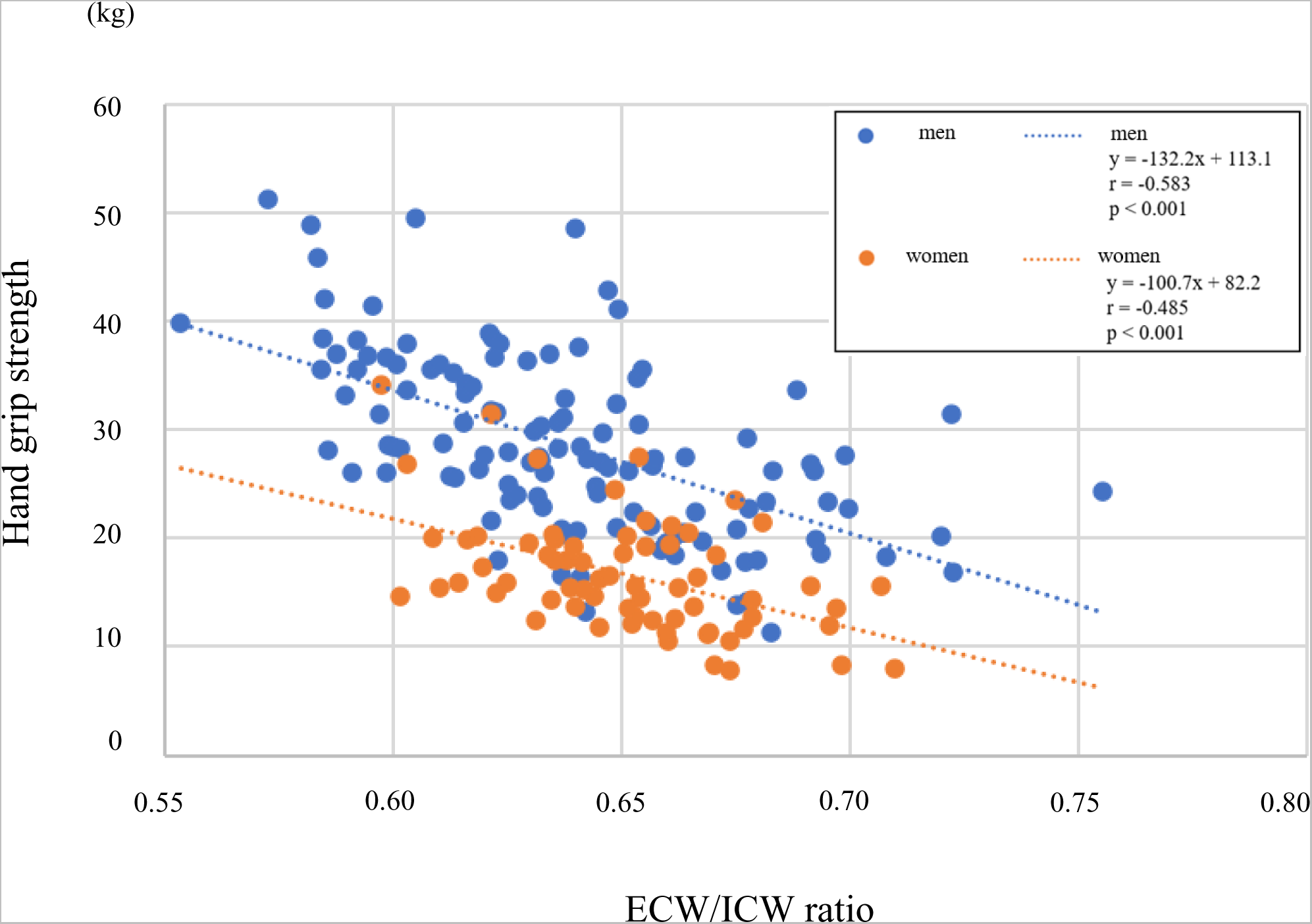
Relationship between high ECW/ICW ratio and handgrip strength for men and women. ICW, intracellular water; ECW, extracellular water.

Figure 3 shows the results of the multivariate logistic regression analysis for predicting low muscle strength as defined by the Asian Working Group for Sarcopenia. We used the three models to examine the factors associated with low muscle strength. In Models 1 and 2, we included the factors that were significant in the univariate analysis (p < 0.05) and the ECW/ICW ratio as a continuous and categorical variable, respectively. We found that a high ECW/ICW ratio and low SMI were independently associated with low muscle strength in both models (p < 0.05). In Model 3, we replaced the ECW/ICW ratio with ECW and ICW separately. We found that both an increasing ECW and decreasing ICW were independently associated with low muscle strength (p < 0.05).

**Figure 3.**
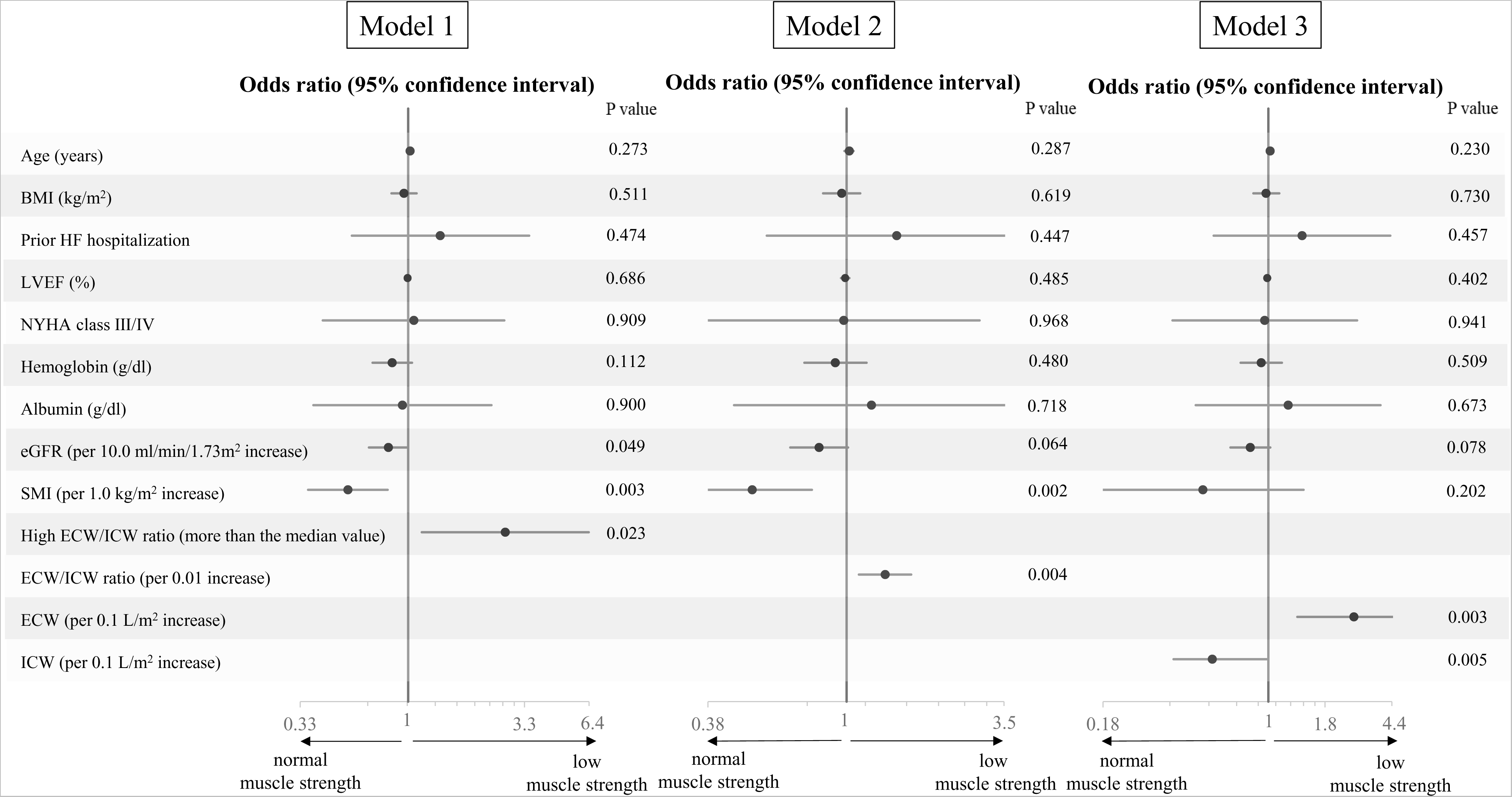
Multivariate logistic regression analysis for predicting low handgrip strength. Models 1 and 2 included the factors that were significant in the univariate analysis and the ECW/ICW ratio as a continuous and categorical variable, respectively. Model 3 included ECW and ICW instead of the ECW/ICW ratio. BMI, body mass index; HF, heart failure; LVEF, left ventricular ejection fraction; NYHA, New York Heart Association; eGFR, estimated glomerular filtration rate; SMI, skeletal muscle index; ICW, intracellular water; ECW, extracellular water.

Figure 4 depicts the relationship between skeletal muscle structure and handgrip strength in a three-dimensional diagram using thin plate smoothing splines. An increasing ECW and/or decreasing ICW was associated with an increasing ECW/ICW ratio and reduced handgrip strength (p < 0.05) (Figure 4A, B). Handgrip strength decreased with a decreasing SMI, and an increasing ECW/ICW ratio further decreased handgrip strength regardless of the SMI value (p < 0.05) (Figure 4C). Therefore, a high ECW/ICW ratio (i.e., an increasing ECW and/or decreasing ICW) promotes muscle weakness regardless of skeletal muscle mass.

**Figure 4.**
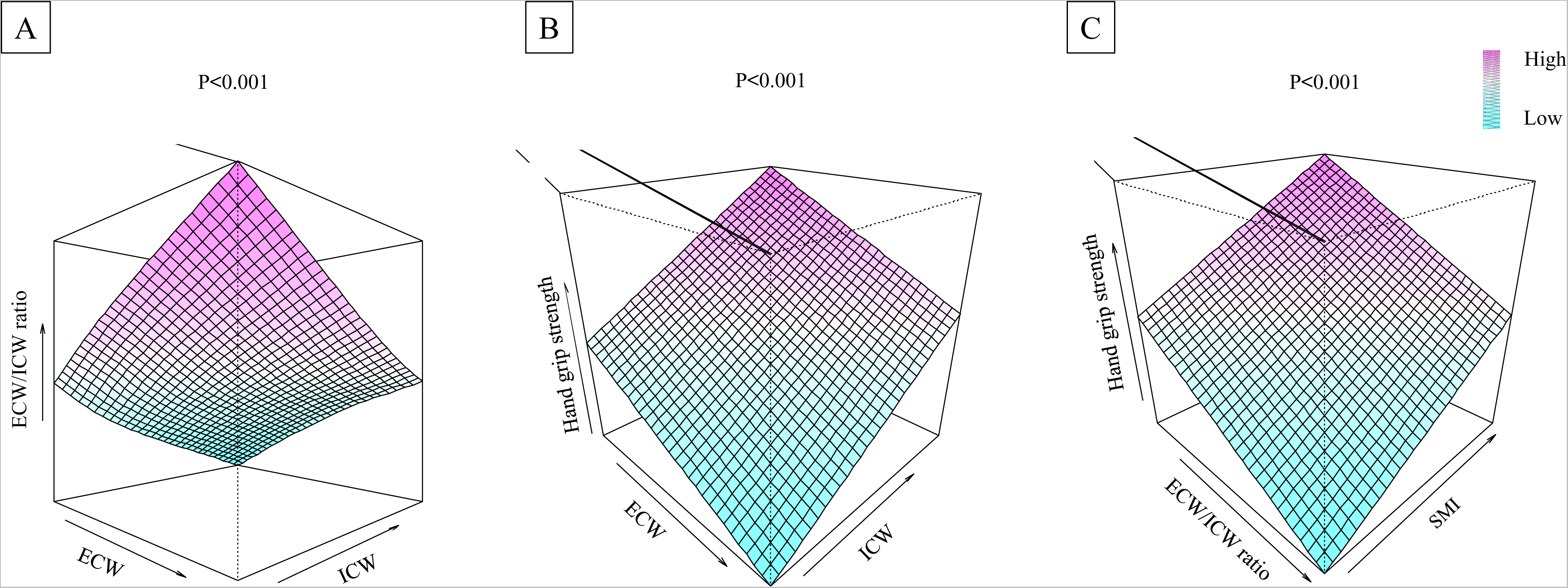
Relationship between skeletal muscle structure and muscle strength in a three-dimensional diagram using thin plate smoothing splines. (A) Relationship between ECW, ICW, and the ECW/ICW ratio. (B) Relationship between ECW, ICW, and handgrip strength. (C) Relationship between ECW/ICW ratio, SMI, and handgrip strength. ICW, intracellular water; ECW, extracellular water; SMI, skeletal muscle index.

## Discussion

This study demonstrated that a high ECW/ICW ratio of skeletal muscle was independently associated with muscle weakness regardless of skeletal muscle mass in patients with HF.

Patients with HF have structural and functional abnormalities of skeletal muscle as well as loss of muscle mass.^3,18^ In this study, a higher ECW/ICW ratio was associated with lower handgrip strength even with the same muscle mass (Figure 4). Thus, the ECW/ICW ratio provides additional information about how the skeletal muscle structure is related to muscle strength apart from muscle mass. A recent study also showed that a high ECW/ICW ratio was associated with decreased muscle strength in patients with HF patients, but the sample size was small (n = 51).^19^ The present study expanded on this finding in a larger sample (n = 190) by demonstrating the relationship of handgrip strength with the ECW and ICW separately.

The mechanisms underlying how a high ECW/ICW ratio is related to muscle weakness in patients with HF are as follows. A high ECW/ICW ratio reflects a decrease in ICW and/or an increase in ECW (Figure 4). A decrease in ICW and an increase in ECW were independently associated with muscle weakness (Figure 3). Thus, both a decreasing ICW and an increasing ECW contribute to the mechanism by which a high ECW/ICW ratio is associated with muscle weakness. ICW reflects the volume of muscle cells. Aging and malnutrition reportedly lead to loss and atrophy of muscle cells, resulting in decreasing ICW and muscle strength.^20^ When muscle cells shrink, more space is created between muscle cells (graphical abstract). This leads to lower ICW and a relative increase in ECW, resulting in a high ECW/ICW ratio.^10^ If muscle volume remained the same, muscles with higher ECW/ICW ratios would have relatively fewer myocytes and lower muscle strength.^21^ We found that a high ECW/ICW ratio was associated with older age and malnutrition, characterized by a low body mass index and low albumin level (Table 1). Therefore, aging and malnutrition may lead to loss and/or atrophy of muscle cells and result in low ICW, a high ECW/ICW ratio, and muscle weakness. The enlarged intercellular spaces directly impair the diffusion of oxygen from capillaries to myocytes, resulting in muscle weakness due to insufficient oxygen supply.^3,22^ This study showed that increasing ECW was associated with muscle weakness independently of decreasing ICW. Thus, even if the volume of myocytes was the same, muscles with a higher ECW/ICW ratio would have more impaired muscle strength due to enlarged intercellular spaces and impaired oxygen diffusion. All patients in the present study had no clinical signs of congestion at the time of data collection. However, we found that a high ECW/ICW ratio was associated with a high B-type natriuretic peptide level, low renal function, and hypoalbuminemia (Table 1). These conditions may be associated with muscle edema due to subclinical fluid retention, resulting in an increasing ECW and ECW/ICW ratio.^6,22^ Thus, the expansion of the intercellular space due to fluid retention and the subsequent increase in the ECW and ECW/ICW ratio may be associated with muscle weakness.

This study had three main limitations. First, it was a single-center, retrospective, observational study. Second, the sample size was relatively small. Third, the data did not allow us to conclude whether a high ECW/ICW ratio is a cause or effect of muscle weakness. Although the speculated pathogenesis suggests that interventions to increase ICW and decrease ECW (i.e., overcoming malnutrition and reducing muscle edema) may help to recover the muscle strength in patients with HF who have a high ECW/ICW ratio and muscle weakness, future interventional studies are necessary to confirm this hypothesis.

In conclusion, a high ECW/ICW ratio of skeletal muscle was associated with loss of muscle strength independently of skeletal muscle mass in patients with HF. Future interventional studies are necessary to establish treatment strategies based on the ECW/ICW ratio.

## Acknowledgment

The authors would like to thank Angela Morben, DVM, ELS, from Edanz (https://jp.edanz.com/ac) for editing a draft of this manuscript.

## Source of Funding

This study was supported by a grant from the Japan Society for the Promotion of Science (JSPS KAKENHI Grant No. 19K08582).

## Disclosures

KY received research grants from Abbott Co., Ltd.; Otsuka Pharmaceutical Co., Ltd.; Biotronik Japan Inc.; Japan Lifeline Co., Ltd.; and Fukuda Denshi. KY also received speakers’ bureau fees from Otsuka Pharmaceutical Co., Ltd.; Novartis; and Daiichi-Sankyo Co., Ltd.

## Data availability statement

The research data will not be shared.

## Nonstandard Abbreviations and Acronyms

ECW: extracellular water
ICW: intracellular water

## Notes

### Author Declarations

The research ethics committee of Tottori University approved the study. (No.2418)

